# Ruxolitinib versus Dexamethasone in Hospitalized Adults with Covid-19: multicenter matched-controlled study

**DOI:** 10.1101/2021.04.20.21255662

**Authors:** O.V. Stanevich, D.S. Fomina, I.G. Bakulin, Sh. I. Galeev, E.A. Bakin, V.A. Belash, A.N. Kulikov, A.A. Lebedeva, D.A. Lioznov, Yu S. Polushin, I.V. Shlyk, E.A. Vorobyev, S.V. Vorobyeva, T.V. Surovceva, N.V. Bakulina, M.A. Lysenko, I.S. Moiseev

## Abstract

**Background:** Several anti-cytokine therapies were tested in the randomized trials in hospitalized patients with severe acute respiratory syndrome coronavirus 2 infection (COVID-19). Both janus kinase (JAK) inhibitor, baricitinib, and dexamethasone demonstrated the reduction of mortality. In this matched control study we compared dexamethasone to another JAK inhibitor, ruxolitinib.

**Methods:** The study included 146 hospitalized patients with COVID-19 and oxygen support requirement. The control group was selected 1:1 from 1355 dexamethasone-treated patients and was matched by 29 clinical and laboratory parameters predicting survival.

**Results:** Ruxolitinib treatment in the general cohort of patients was associated with equivalent to dexamethasone mortality rate: 9,6% (95% CI 4,6-14,6%) vs 13,0% (95% CI 7,5-18,5%, superiority p=0.35, non-inferiority p=0.0137), respectively. Time to discharge without oxygen support requirement was also not different between these groups: 13 vs 11 days (p=0.13). Subgroup analysis without adjustment for multiple comparisons demonstrated reduced mortality in ruxolitnib-treated patients with febrile fever (OR 0.33, 95%CI 0.11-1.00). Except higher incidence of grade 1 thrombocytopenia (37% vs 23%, p=0.042), ruxolitinib therapy was associated with better safety profile due to reduced rate of severe cardiovascular adverse events (6.8% vs 15%, p=0.025).

**Conclusions:** Ruxolitinib may be an alternative anti-cytokine therapy with comparable efficacy in patients with potential risks of steroid administration. Patients with febrile fever at admission may benefit from ruxolitinib administration.

**Funding:** Ruxolitinib was obtained from Novartis through Managed Access Program (MAP).

## Introduction

It is now established that excessive production of cytokines is an important part of pathogenesis during severe acute respiratory syndrome coronavirus 2 (SARS-CoV-2) infection (COVID-19) [1,2]. Usually patients present with elevation of multiple cytokines [3,4]. Clinical consequence of increased cytokine production is the development of hyperinflammatory syndrome (HIS), also called cytokine-release syndrome, or macrophage-activation syndrome, or secondary haemophagocytic lymphohistiocytosis. Symptoms of HIS usually include high fever, disseminated intravascular coagulation (DIC), acute respiratory distress syndrome (ARDS), encephalopathy, abnormal liver function tests, acute kidney injury, lymphopenia, low platelet count [2,5].

Based on these observations of HIS various types of anti-cytokine therapies were evaluated in COVID-19 patients, including anti-interleukin-6 (IL-6)6, anti-interleukin-1 (IL-1)7 and anti-granulocyte-monocyte colony-stimulating factor (GM-CSF)[8]. Despite faster resolution of symptoms was reported in these studies so far only dexamethasone [9] and baricitinib [10], a Janus kinase (JAK) 1 and 2 inhibitor, demonstrated improved survival in patients hospitalized for COVID-19 compared to the standard of care.

Another JAK 1/2 inhibitor, ruxolitinib, also demonstrated effective anti-cytokine properties in myelofibrosis [11], graft-versus host disease (GVHD) [12] and secondary haemophagocytic lymphohistiocytosis [13]. Also it was reported to improve COVID-19 clinical course in several small patient serieses [14,15]. Recently the press release indicated that randomized RUXCOVID study did not meet its primary endpoint of reduced cumulative incidence of death mechanical ventilation or ICU care compared to the standard of care [16]. Since the ‘standard’ of care for COVID-19 is constantly changing and, unlike the early studies, now the majority of severe patients do receive some form of anti-cytokine therapy, we compared ruxolitinib to the most common immunosuppressive treatment, dexamethasone, in the multicenter matched-control study.

## Methods

### Patients

The study was conducted in four large specialized hospitals for COVID-19 infection in Saint Petersburg and Moscow. The active treatment arm included 146 patients from the Ruxolitinib Managed Access Program (MAP) for Patients Diagnosed With Severe/Very Severe COVID-19 Illness (clinicaltrials.gov NCT04337359, CINC424A2001M). The inclusion criteria in this analysis were polymerase chain reaction (PCR)-confirmed case of COVID-19 infection and 5-6 score on the Ordinal scale^17^. Patients with scores 4 and less were excluded because these patients generally receive outpatient care in Russian Federation and the sample of hospitalized patients is not representative of the general population. Patients with score 7 and higher were excluded because there is limited data on pharmacokinetics of ruxolitinib after administration of crashed tablets via gastric tube. The only exclusion criteria in the analysis was terminal oncological illness on palliative care. All consecutive patients meeting the inclusion and exclusion criteria were included in the analysis.

COVID-19 infection was confirmed by both the nasopharyngeal and oropharyngeal swab with RealBest RNA SARS-CoV-2 test kit for real time PCR (Vector-Best, Novosibirsk, Russian Federation). Patients were followed up until death or discharge. Local ethical committees approved the participation of patients in the Managed Access Program and all patients signed informed consent for the treatment. Ruxolitinib was administered in doses 5-10 mg bid. Median dose was 0.125 was mg/kg/day. Dose distribution per kilogram of patient weight is presented in supplementary figure S1. Ruxolitinib was administered until oxygen support withdrawal and discontinued abruptly. Computer tomography (CT) severity grade was assessed using Russian Ministry of Health recommendations. In brief the grading system is based on the percent of effected lung tissue: grade 1 (<25%), grade 2 (25-50%), grade 3 (51-75%), grade 4 (>75%)[17].

The control group was selected from 1355 contemporary hospital-stratified patients receiving dexamethasone 16-24 mg of dexamethasone daily for 5-10 days based on the symptom duration.

### Outcome measures, group matching and statistical analysis

The following major clinical and laboratory disease features were used for selection of patients for the control group: age, gender, body mass index (BMI), SpO2 wiithout oxygen support, C-reactive protein (CRP), additional anti-cytokine therapy, pre-existing diabetes and absolute lymphocyte count. Additional parameters assessed during matching were serum creatinine, serum glucose level, white blood cell (WBC) count, potassium, sodium, hemoglobin, neutrophil count, platelet count, pre-existing cardiovascular, respiratory, liver disorders, tuberculosis and oncological disease requiring active treatment. CT grade was not included in the matching parameters because it was not yet validated for mortality. Only pre-treatment laboratory measures were used for matching. The propensity score matching was based on all of these parameters and facilitated selection of the control group with 1:1 ratio. The balance of covariates before and after matching is given in supplementary figure S2. Contingency table for mortality before and after matching is available in supplementary table S1.

Moreover, to confirm the significance of the parameters selected for the matching in the ruxolitinib group we conducted a cluster analysis based on selected matching variables. The analysis effectively predicted survival in the study group. The survival in the three selected clusters was 85% vs 98% vs 87%, p=0.0087 (Supplementary figure S4, supplementary tables S2). Although the difference in survival between cluster 1 and 3 was modest, however the algorithm effectively selected in the cluster 2 patients with significant risk of death without prominent SpO2 decrease upon admission. The verification of the control group selection procedure demonstrated that there was a fair matching within each of the clusters (supplementary figure S5, supplementary tables S4).

WHO recommendations for selection of outcomes were used to define the outcome of the study^19^. Cumulative incidences of death and discharge as competing risks were selected as the primary outcome of the study based on the following considerations: 1) universal healthcare system in Russian Federation excludes discharges for economic reasons, which is the main concern about discharge endpoint; 2) we were not limited in the follow up time of intubated or discharged patients, so either one or another outcome did occur within the scope of the study; 3) time to PCR-negativity was not appropriate endpoint in this study because some asymptomatic patients were discharged before PCR-negativity. The differences in clinical measures, laboratory parameters and toxicity between study groups were assessed with non-parameteric tests: Chi-square and Mann-Whitney tests according to the data type. Cumulative incidence analysis and Gray test were used to compare mortality and discharge rates between dexamethasone and ruxolitinib. Farrington-Manning test with 10% margin was used to demonstrate non-inferior discharge rates. Logistic regression regression with mortality as an outcome was used for the subgroup analysis. CRP threshold for subgroup analysis was selected from the set of dexamethasone patients not included in the main analysis. In this group of patients receiver operating curve analysis was performed targeting highest average sensitivity and specificity for in-hospital mortality. The resulting threshold level was approximately 100 mg/L (Supplementary figure S4). All analyses were performed using R statistics, version 4.0.4. The packages used for the analysis are listed in the supplementary Table S5.

## Results

Analyzed groups of patients with ruxolitinib (N=146) and dexamethasone (N=146) treatments were well matched in their clinical and laboratory parameters (Table 1). Cumulative incidence of mortality was 9.6% in the ruxolitinib group and 13.0% in the dexamethasone group (superiority p=0.35, non-inferiority p=0.0137, Figure 1). Median time to discharge was 13 vs 11 days for these groups, respectively (p=0.13). The subgroup analysis of mortality without adjustment for multiple comparisons demonstrated that dexamethasone was not superior to ruxolitinib in any of the subgroups tested. However, ruxolitinib administration was associated with borderline mortality reduction in patients with high persistent fever (OR 0.33, 95%CI 0.11-1.00). Also, surprisingly, improved survival was observed not in patients with cardiovascular disease who were expected to tolerate steroids worse than ruxolitinib, but in patients without cardiovascular disease (OR 0.23, 95%CI 0.06-0.88, Figure 2). None of the other complete blood count parameters (Supplementary figure S6), biochemistry parameters (Supplementary figure S7), demographic parameters (Supplementary figure S8) or CT stage (Supplementary table S6) were predisposing to better survival with either of the anti-cytokine therapies.

**Table 1.** Characteristics of patients treated with ruxolitinib and matched control group treated with dexamethasone.

| **Names** | **Reference arm**<br>**(n = 146)** | **Treatment arm**<br>**(n = 146)** | **p-value** |
| --- | --- | --- | --- |
| Age, years, mean±SD | 58±12.9 | 58.1±13.3 | 0,93 |
| Gender | F: 64 (43.8%)<br>M: 82 (56.2%) | F: 66 (45.2%)<br>M: 80 (54.8%) | 0,81 |
| Body mass index, mean±SD | 30.7±13.4 | 30.4±5.6 | 0,81 |
| Day of illness | 7.4±4.3 | 7.5±4.3 | 0,87 |
| Admission creatinine, mmol/L | 0.1±0 | 0.1±0 | 0,67 |
| Admission CRP, mg/L | 99.5±84.7 | 97.4±71.9 | 0,82 |
| Maximal CRP, mg/L | 105.2±80.6 | 101.1±86.4 | 0,68 |
| Admission ferritin, ng/mL | 646±538.3 | 617±403.8 | 0,6 |
| Maximal glucose, mmol/L | 7.7±2.7 | 7.6±2.9 | 0,71 |
| Maximal temperature, °C | 38.1±0.8 | 38.1±0.9 | 0,77 |
| Mean lymphocytes, 10<sup>9</sup>/L | 1.2±0.5 | 1.3±0.8 | 0,074 |
| Mean monocytes, 10<sup>9</sup>/L | 0.4±0.2 | 0.5±0.3 | 0,41 |
| Mean neutrophils, 10<sup>9</sup>/L | 5.3±3.8 | 5.1±3.1 | 0,72 |
| Mean potassium, mmol/L | 4±0.4 | 4±0.5 | 0,95 |
| Mean sodium, mmol/L | 138.7±3.4 | 138.6±3.8 | 0,83 |
| Mean WBC, 10<sup>9</sup>/L | 6.9±3.8 | 6.8±3.2 | 0,79 |
| Admission hemoglobin, g/L | 133.5±18.7 | 134.4±19 | 0,69 |
| Minimal platelets, 10<sup>9</sup>/L | 202.2±68.9 | 201.1±64.2 | 0,89 |
| Admission SpO<sub>2</sub>, % | 93.7±2.3 | 93.6±3.2 | 0,74 |
| Minimal SpO<sub>2</sub>, % | 93.8±3.3 | 93.5±4.6 | 0,43 |
| Cardiovascular disease | Yes: 75 (51.4%)<br>No: 71 (48.6%) | Yes: 73 (50%)<br>No: 73 (50%) | 0,82 |
| Diabetes | Yes: 33 (22.6%)<br>No: 113 (77.4%) | Yes: 29 (19.9%)<br>No: 117 (80.1%) | 0,57 |
| HIV | Yes: 2 (1.4%)<br>No: 144 (98.6%) | Yes: 1 (0.7%)<br>No: 145 (99.3%) | 0,56 |
| Chronic liver disease | Yes: 5 (3.4%)<br>No: 141 (96.6%) | Yes: 5 (3.4%)<br>No: 141 (96.6%) | 1 |
| Oncological disease not in remission | Yes: 7 (4.8%)<br>No: 139 (95.2%) | Yes: 9 (6.2%)<br>No: 137 (93.8%) | 0,61 |
| Chronic respiratory disease | Yes: 11 (7.5%)<br>No: 135 (92.5%) | Yes: 17 (11.6%)<br>No: 129 (88.4%) | 0,23 |
| Tuberculosis | Yes: 4 (2.7%)<br>No: 142 (97.3%) | Yes: 3 (2.1%)<br>No: 143 (97.9%) | 0,7 |
| Severity of lung injury based on computed tomography | CT-1: 16<br>CT-2: 83<br>CT-3: 47 | CT-1: 20<br>CT-2: 97<br>CT-3: 29 | 0,058 |

All laboratory values include levels before either ruxolitinib or dexamethasone administration. Laboratory values are presented as mean±standard deviation. CRP=C-reactive protein, HIV=human immunodeficiency virus infection.

**Figure 1.**
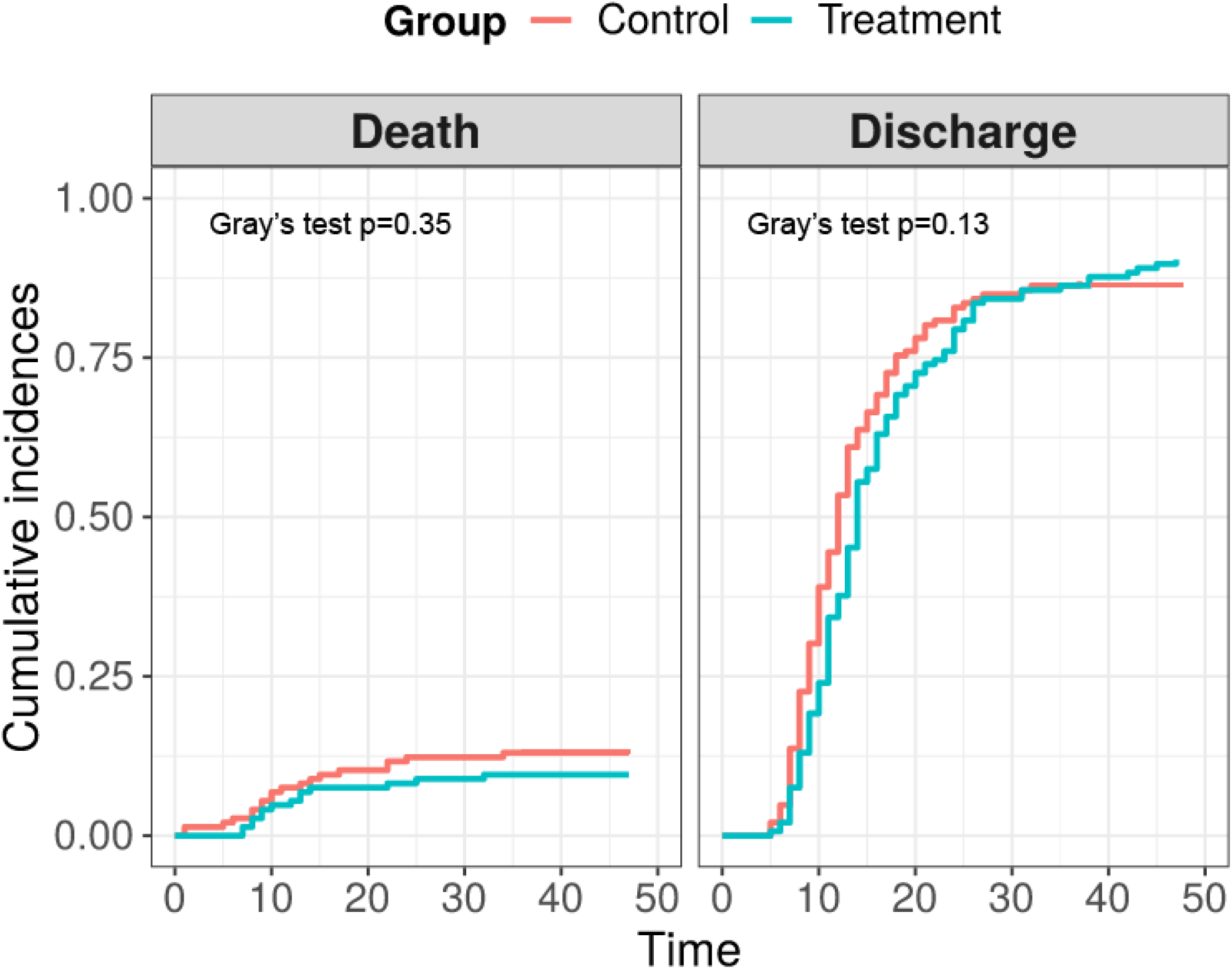
Cumulative incidence of death and discharge in the dexamethasone (control) and ruxolitinib groups (treatment).

**Figure 2.**
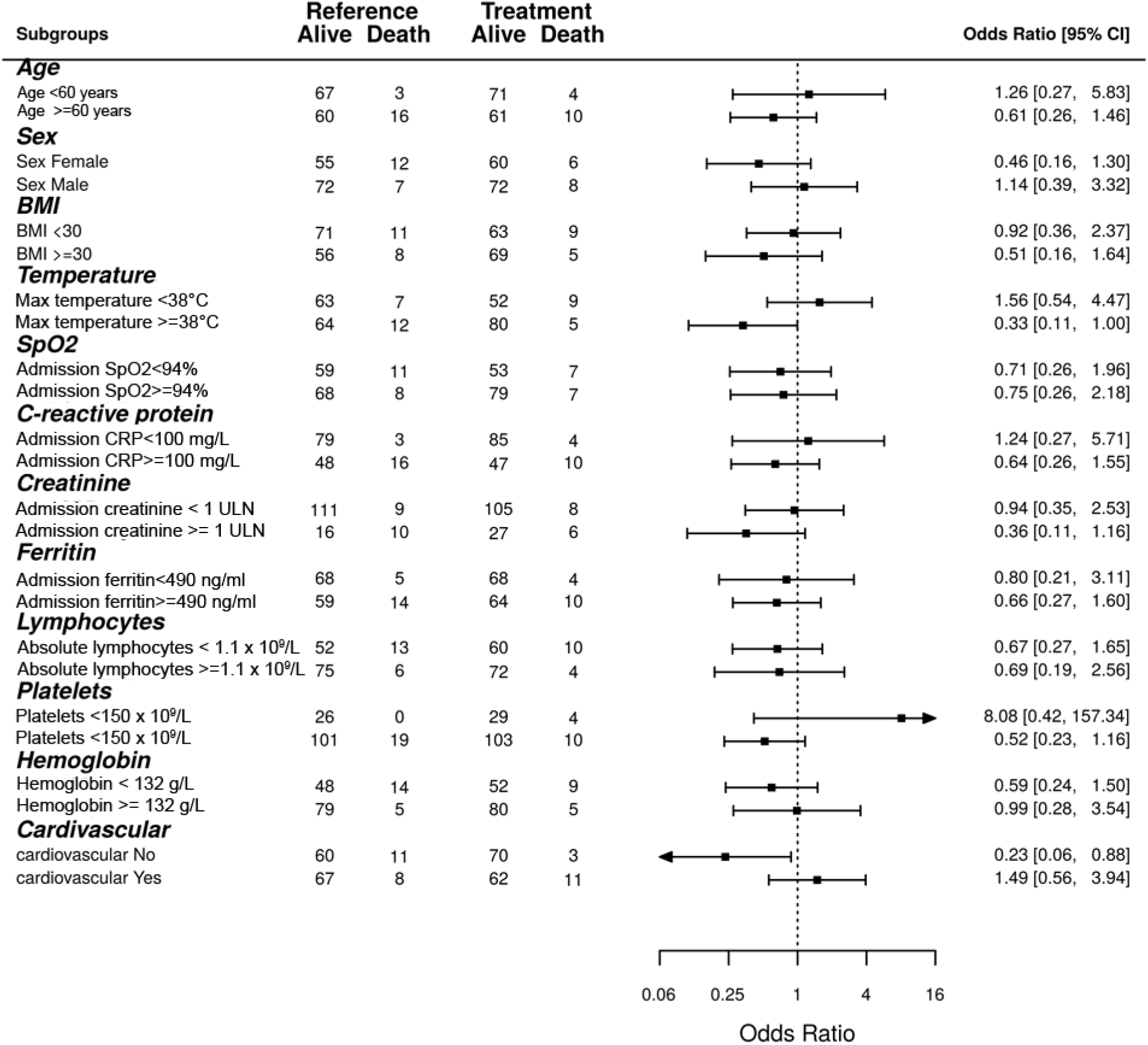
Forrest plot with subgroup analysis of mortality according to the clinical and laboratory characteristics. Bars to the left of the reference line indicate superiority of ruxolitinib, to the right – superiority of dexamethasone. ULN=upper limit of normal. Max=maximal value documented in-hospital before anti-cytokine therapy. The cut off levels of absolute lymphocytes and hemoglobin represent local normal reference ranges.

The analysis of complications demonstrated almost equal distribution of hematological adverse events between groups (Supplementary table S7). Only grade 1 thrombocytopenia was observed more often after ruxolitinib (37% vs 23%, p=0.042). The incidence of grade 2-4 thrombocytopenia was not different between the groups (4.1% vs 3.4%). However, fewer severe cardiovascular adverse events were observed in the ruxolitinib group (6.8% vs 15%, p=0.025), including reduction in the incidence of pulmonary embolism (2.0% vs 7.5%, p=0.028) and trend to decreased incidence of acute myocardial infarction (4.8% vs 10.3%, p=0.076). Incidence of deep vein thrombosis was not different between two arms (1.4% vs 1.4%, p=1.0). Overall incidence of secondary severe infectious adverse events was not different between groups (7.5% vs 9.6%, p=0.53). The incidence of nosocomial bacterial pneumonias (3.4% vs 4.1, p=0.76) and sepsis (4.1% vs 6.1%, p=0.43) was also not different in the ruxolitinib and dexamethasone groups, respectively. The list of rare adverse events is presented in the supplementary table S8.

## Discussion

Although, World Health Organization (WHO) does not recommend routine use of immunosuppressive therapies [20], dexamethasone [9] and JAK inhibitor baricitinib [10] were the one of the few agents that demonstrated improvement of survival in large populations of patients with severe COVID-19 infection. In our study in the general group there was no significant differences between the evaluated agents in prevention of mortality, however dexamethasone administration was associated with faster hospital discharge. Despite absence of survival advantage in the general population, during the matching process we observed that COVID-19 is a very heterogeneous disease. This might be the reason for the failure of several randomized trials [6,21]. Thus from an attempts to find a universal cure for all COVID-19 patients it is important to formulate which patient population is the candidate for immunosuppressive therapy and what kind of anti-cytokine therapy should be applied in different clinical situations.

It is clear that at least in part this heterogeneity in COVID-19 infection comes from HIS clinical presentation. Before COVID-19 pandemic, it was evident that each etiology of HIS, or CRS, is associated with unique clinical features. CRS after chimeric antigen receptor T cells is characterized by neurotoxicity and hypotension [22]. In rheumatic diseases most common clinical features are hepatomegaly, splenomegaly, polyserositis and cytopenias [23]. In sepsis and hematological malignancies it is multiorgan failure with overproduction of serum ferritin [24,25]. HIS in severe COVID-19 is another distinct entity with mild to moderate organ damage, but severe endothelial dysfunction, ARDS, hypercoagulation state and microangiopathy-like cytopenias [2,26,27]. Renal pathological findings in COVID-19 HIS are also quite unique and are characterized by tubular damage with abnormal sodium reabsorption, microangiopathy with hypoperfusion and glomerulopathy [28].

We observed that ruxolitinib improved survival in patients with persistent fever (>38°C), which is one of the key features of COVID-19 HIS [2]. Besides fever other clinical features of HIS, like acute renal injury or high CRP did not reach statistical significance, since they were observed in the minor subsets of patients, so we were unable with the current study size to formulate the exact HIS feature where patients can benefit from ruxolitinib. However, the ability of ruxolitinib to control inflammation with endothelial damage is not unexpected. It was recently approved for steroid-refractory GVHD after allogeneic stem cell transplantation [12]. Steroid-refractory GVHD is another hyperinflammatory condition with severe endothelial dysfunction [29,30]. The unique role of ruxolitinib in the situation of abnormal inflammation and endothelial dysfunction is confirmed by the fact that it is the first therapy approved for steroid-refractory acute GVHD in 30 years despite multiple academic studies of various immunosuppressive agents [31,32]. Thus, it is unclear if other anti-cytokine therapies will provide similar results to JAK inhibitors.

On the contrary, mortality was comparable between dexamethasone and ruxolitinib across all subgroups of pulmonary severity, including admission SpO2 and computer tomography (CT) stage. Absence of difference can be explained by complex pathogenesis of lung injury. It was reported that besides HIS-associated interstitial edema and endothelial dysfunction the following events may contribute to the damage: thrombosis of large and small vesels, formation of hyaline membranes, complement-associated injury, impaired surfactant production and direct viral cell injury [33,34,35]. Thus both dexamethasone and ruxolitinib may ameliorate HIS-associated components of lung injury, but not the other.

The major concern about anti-cytokine therapies, which was also stated in the WHO treatment guidelines [20], is the risk of secondary infections and other complications. Hematological toxicity is a known complicaition of ruxolitinib, however it is generally observed in patients with underlying hematological disease [11,12]. In the doses used in this study, no clinically relevant hematological toxicity was observed. Although grade 1 thrombocytopenia was more prevalent after ruxolitinib, in some patients we observed, on the contrarily, resolution of HIS-associated cytopenias during treatment. The other concern about immunosuppressive agents in COVID-19 patients are secondary nosocomial infections. Although the rate of hospital acquired infections vary across countries, in the studied cohort their incidence was comparable to an intensive care of other conditions [36]. The reduced incidence of cardiovascular adverse events is not surprising. One the one hand, COVID-19 infection itself predisposes to venous thromboembolism, and some studies report the incidence of this complication as high as 37% [37]. On the other hand, the prothrombotic effects of steroids are known for a long time [38]. High incidence of myocardial infarction and venous thromboembolism emerged despite direct anticoagoalation therapy in all patients. One of the possible explanations is the microangiopathic origin of the thrombosis during COVID-19 infection, where anti-complement therapy might be a more effective way to prevent thrombotic events [39]. Our results confirm a favorable safety profile seen with another JAK inhibitor, baricitinib, in the randomized trial [10].

In conclusion, we demonstrated that in the general cohort of patients with severe COVID-19 infection ruxolitinib was equivalent to dexamethasone in preventing in-hospital mortality. Survival benefit was observed in the ruxolitinib group in patients with high fever and without additional cardiovascular co-morbidity. Recently announced results of randomized RUXCOVID trial demonstrated no difference in the primary composite endpoint [16], but it is unclear how this study was balanced in terms of HIS symptoms and COVID-19 severity based on WHO scoring system.

## Data Availability

No applicable

## Acknowledgements

We thank Novartis for providing access to ruxolitinib through Managed Access Program.

## Conflict-of-interest disclosure

I.S.M. had received travel grants from MSD, Novartis, Pfizer, Celgene, Takeda, BMS, consulting fees from Novartis and Celgene, lecturer fees from Novartis, Takeda and Johnson&Johnson.

## Notes

### Competing Interest Statement

The authors have declared no competing interest.

### Clinical Trial

NCT04337359

