## Supplementary figure S1-S7, Supplementary table S1-S8 for "Ruxolitinib versus Dexamethasone in Hospitalized Adults with Covid-19: multicenter matched-controlled study"

Supplementary figure S1. Dose distribution in mg/kg of patient weight.

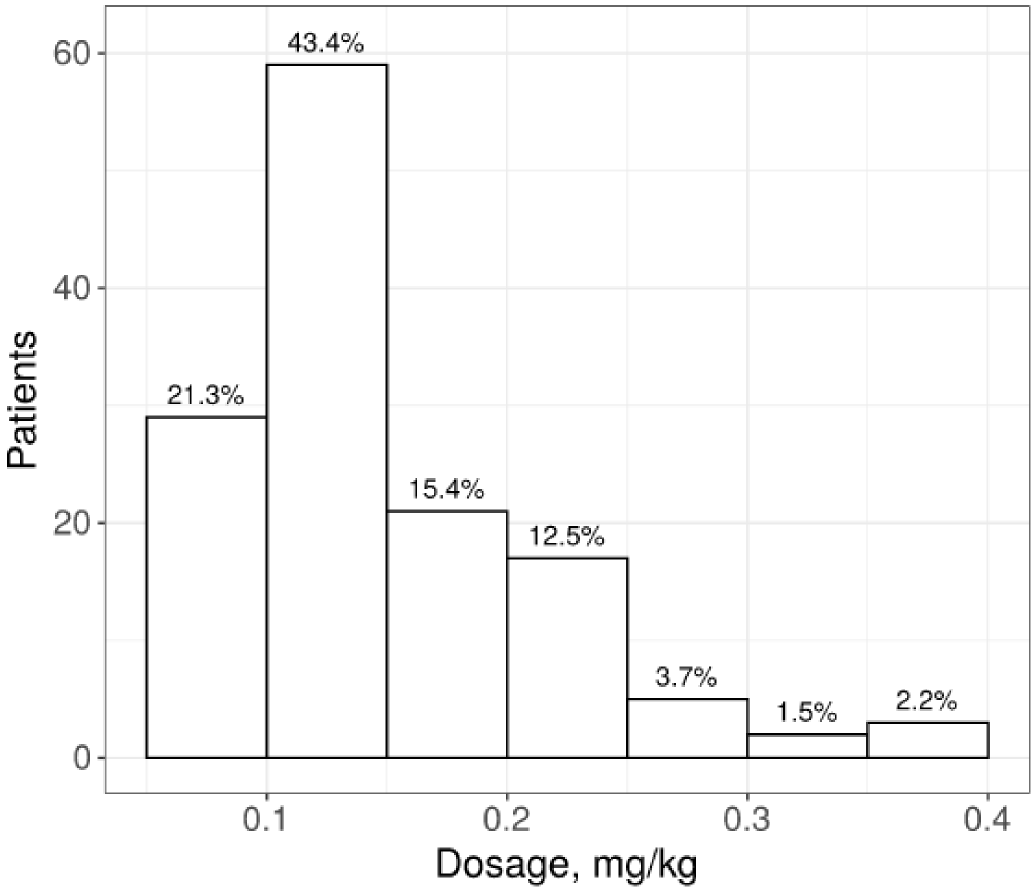

Supplementary figure S2. Covariate balance plot.

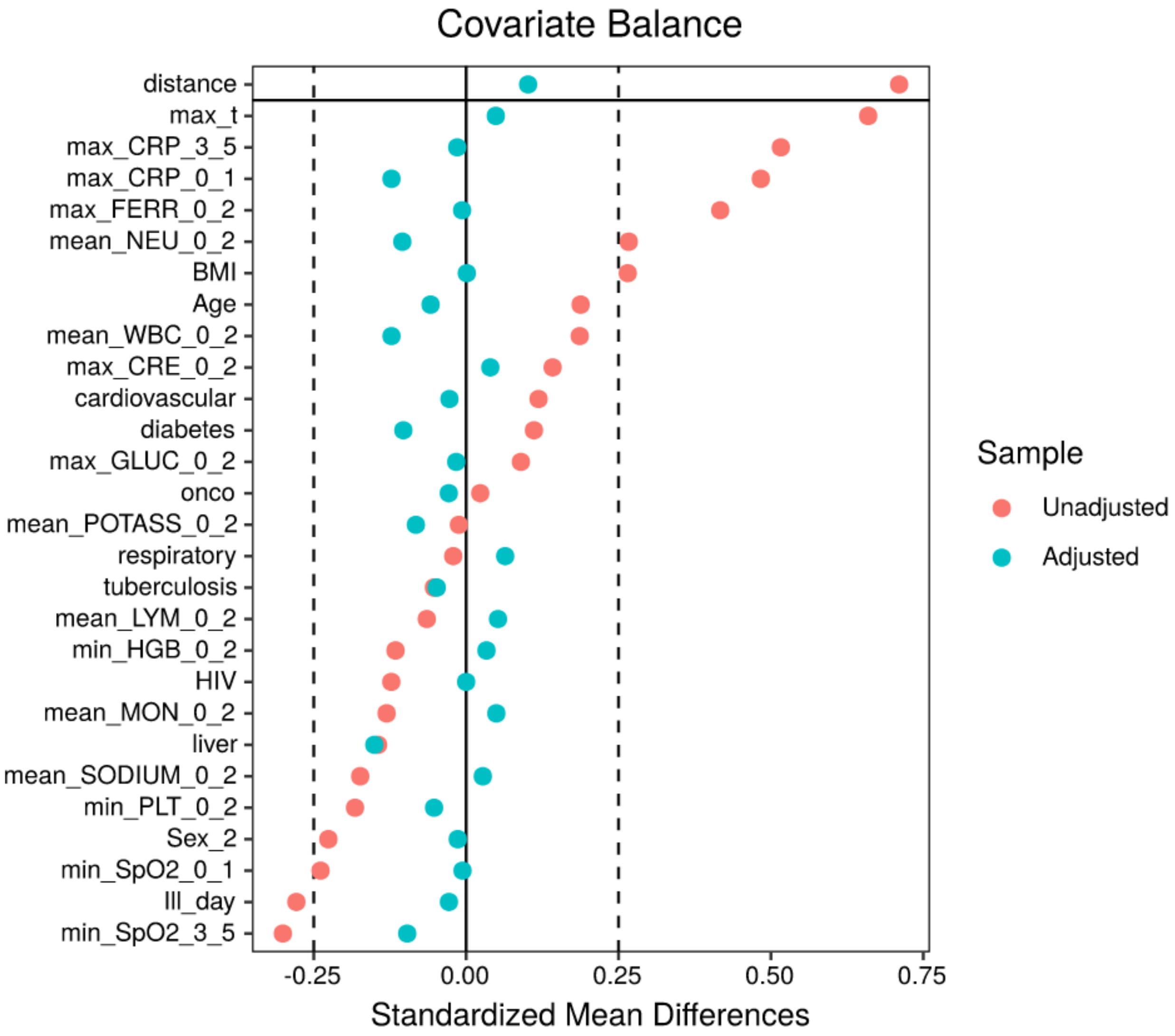


Red dots represent the standardized mean differences between the whole dexamethasone cohort (N=1226) and ruxolitinib group (N=146), while blue dots represent the difference between ruxolitinib group and 146 selected matched control patients. The vertical dotted lines indicate represent target matching quality of ±0.25. Max_t= maximal body temperature, max_CRP_0_1=admission C-reactive protein level, max_CRP_3_5= maximal C-reactive protein level before study treatment administration, mean_NEU_0_2= mean neutrophil level before study treatment administration, mean_WBC_0_2= mean white blood cell count before study treatment administration, Another_anticyt= use of additional anticytokine therapies during hospitalization, BMI=body mass index, max_ max_CRE_0_2=admission creatinine level before study treatment administration, min_NEU= minimal level of neutrophils before study treatment administration, fibrillacia=atrial febrilation, cardiovascular= any cardiovascular co-morbidity, max_GLUC_0_2=admission serum glucose level before study treatment administration, mean_POTASS_0_2= mean serum potassium level before study treatment administration, min_WBC= minimal white blood cell count before study treatment administration, mean_LYM_0_2= mean lymphocytes before study treatment administration, onco=any oncological disease currently requiring treatment, respiratory= any concurrent chronic respiratory disease, min_HGB_0_2= administration hemoglobin level, min_PLT_0_2= minimal administration platelet level, mean_SODIUM_0_2= minimal sodium level before study treatment administration, liver= any concurrent chronic liver disease, Sex=gender, Ill_day= day of illness at the time of the study intervention, min_SPO2_0_1= minimal SpO2 without oxygen support upon admission, min_SPO2_3_5= minimal SpO2 before study drug administration without oxygen support.

Supplementary table S1. Contingency table for group outcomes.

| Experiment | Group | Discharge | Death | OR, 95% CI | *p*-value |
|---|---|---|---|---|---|
| Before matching | Treatment arm | 134 | 14 | 1.95, [1.07, 3.53] | 0.036 |
| | Reference arm | 1285 | 70 | | |
| After matching | Treatment arm | 132 | 14 | 0.71, [0.31, 1.57] | 0.46 |
| | Reference arm | 127 | 19 | | |

Supplementary figure S3. Sensitivity and specificity of mortality prediction according to the admission CRP levels.

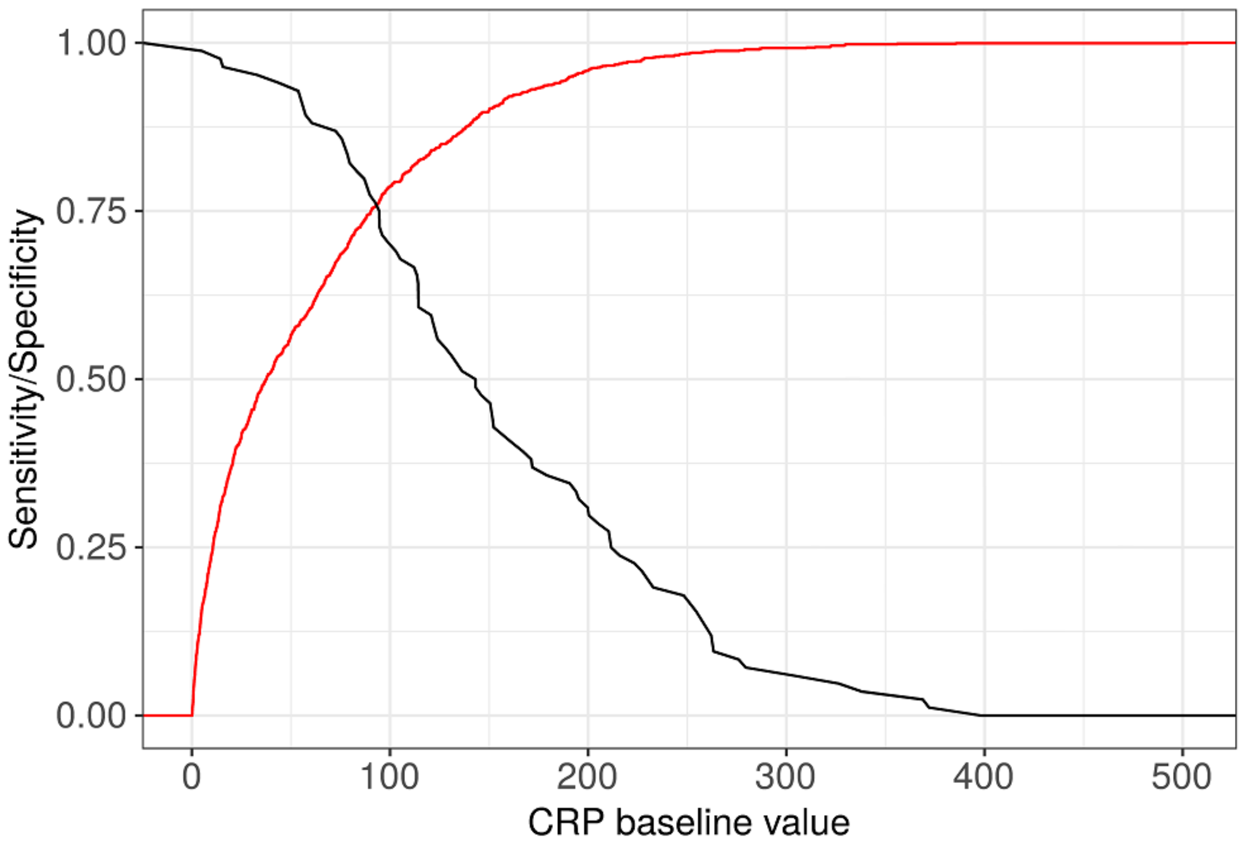


The plot is built on the subset of dexamethasone-treated patients not included in the main analysis (N=1209).

Supplementary figure S4. Cluster analysis of parameters included in the matching procedure in the ruxolitinib group.

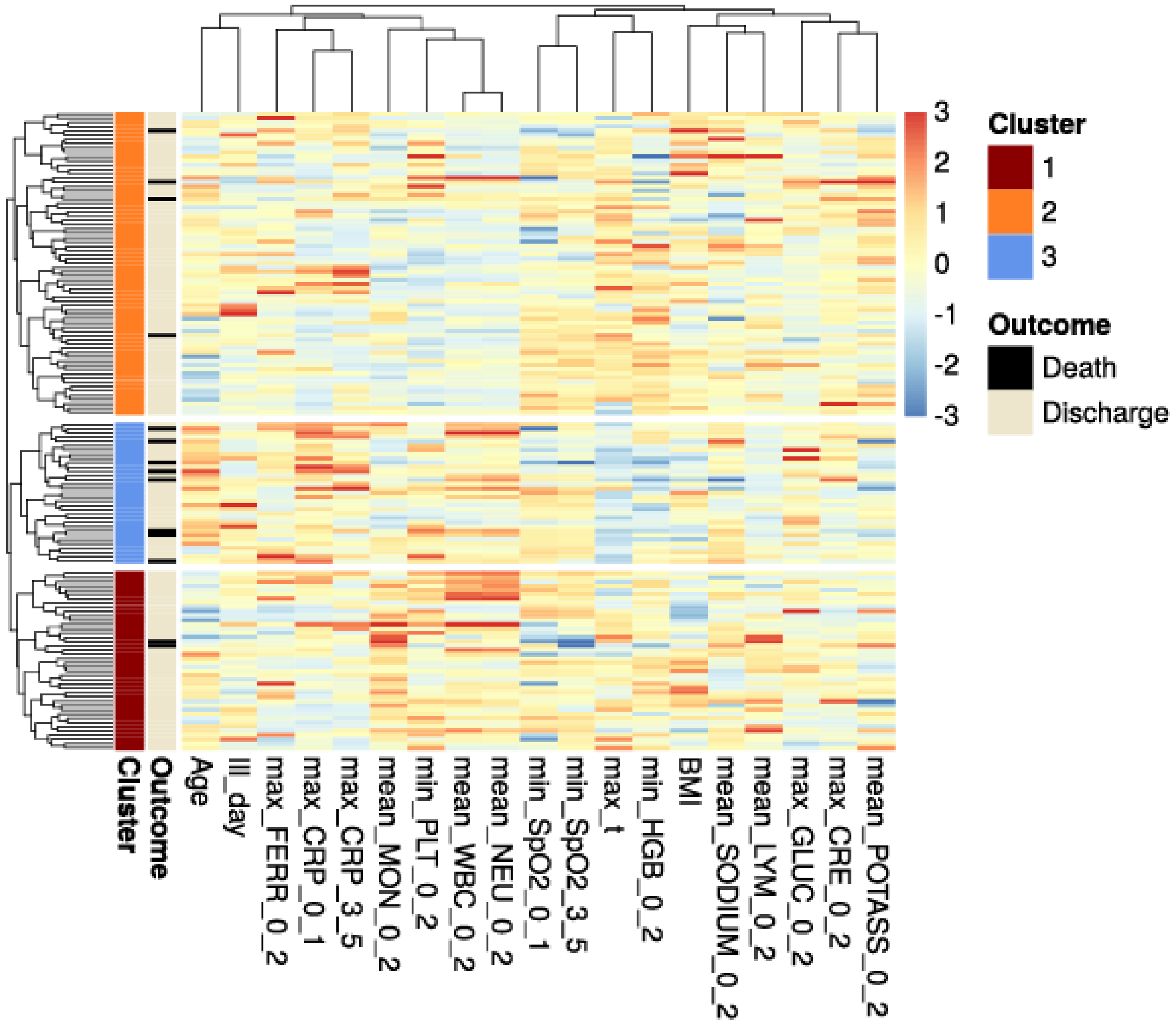


Max_t= maximal body temperature, max_CRP_0_1=admission C-reactive protein level, max_CRP_3_5= maximal C-reactive protein level before study treatment administration, mean_NEU_0_2= mean neutrophil level before study treatment administration, mean_WBC_0_2= mean white blood cell count before study treatment administration, Another_anticyt= use of additional anticytokine therapies during hospitalization, BMI=body mass index, max_ max_CRE_0_2=admission creatinine level before study treatment administration, min_NEU= minimal level of neutrophils before study treatment administration, fibrillacia=atrial febrilation, cardiovascular= any cardiovascular co-morbidity, max_GLUC_0_2=admission serum glucose level before study treatment administration, mean_POTASS_0_2= mean serum potassium level before study treatment administration, min_WBC= minimal white blood cell count before study treatment administration, mean_LYM_0_2= mean lymphocytes before study treatment administration, onco=any oncological disease currently requiring treatment, respiratory= any concurrent chronic respiratory disease, min_HGB_0_2= administration hemoglobin level, min_PLT_0_2= minimal administration platelet level, mean_SODIUM_0_2= minimal sodium level before study treatment administration, liver= any concurrent chronic liver disease, Sex=gender, Ill_day= day of illness at the time of the study intervention, min_SPO2_0_1= minimal SpO2 without oxygen support upon admission, min_SPO2_3_5= minimal SpO2 before study drug administration without oxygen support.

Supplementary table S2. Matching parameters with the clusters.

| Names | Cluster 1 ($n$ = 28) | Cluster 2 ($n$ = 70) | Cluster 3 ($n$ = 47) |
|---|---|---|---|
| Age, years, mean±SD | 62.2±11.5 | 53.2±12.2 | 63.4±13.2 |
| Gender | M: 19 (67.9%)<br>F: 9 (32.1%) | M: 41 (58.6%)<br>F: 29 (41.4%) | M: 20 (42.6%)<br>F: 27 (57.4%) |
| Body mass index | 31.6±5.9 | 29.2±4.9 | 31.1±6.1 |
| Day of illnes | 7±3.4 | 6.6±2.7 | 9±6 |
| Maximal creatinine, mmol/L | 0.1±0 | 0.1±0 | 0.1±0.1 |
| Admission CRP, mg/L | 123.4±82.2 | 61.9±46 | 136.1±73.7 |
| Maximal CRP, mg/L | 93.9±85.3 | 73.7±56.5 | 144.8±105.5 |
| Maximal Ferritin, ng/mL | 727.7±519.4 | 451.9±230.2 | 417.9±131.5 |
| Maximal glucose, mmol/L | 7.4±1.6 | 6.9±1.9 | 8.6±4.2 |
| Maximal temperature, °C | 38±0.9 | 38.3±1 | 37.4±0.9 |
| Mean lymphocytes, $10^9$/L | 1.3±0.5 | 1.5±0.6 | 1±0.9 |
| Minimal lymphocytes, $10^9$/L | 0.9±0.4 | 1±0.5 | 0.6±0.3 |
| Mean monocytes, $10^9$/L | 0.6±0.3 | 0.4±0.2 | 0.4±0.2 |
| Mean neutrophils, $10^9$/L | 8±4.2 | 3.7±1.4 | 5.5±2.9 |
| Minimal neutrophils, $10^9$/L | 4.6±2.2 | 2.6±1.1 | 3.1±1.6 |
| Mean WBC, $10^9$/L | 10±4.3 | 5.4±1.8 | 6.9±2.8 |
| Minimal WBC, $10^9$/L | 6.7±2.7 | 4.5±1.4 | 4.6±1.8 |
| Mean platelets, $10^9$/L | 221.4±66.2 | 181.7±58.4 | 223.6±62 |
| Minimal platelets, $10^9$/L | 183.8±75.6 | 170±53.7 | 185.6±76 |
| Admission hemoglobin, g/L | 140.1±12.3 | 142.3±16.2 | 120.6±16.7 |
| Minimal hemoglobin, g/L | 123.4±19.9 | 129.6±14.9 | 103.8±18 |
| Mean potassium, mmol/L | 3.9±0.6 | 4.1±0.4 | 4±0.5 |
| Mean sodium, mmol/L | 140.6±3.5 | 138±3 | 138.3±4.5 |
| Admission SpO2, % | 92.5±4.7 | 95.8±2 | 95.9±2.2 |
| Minimal SpO2, % | 92.8±3.6 | 94.6±3.3 | 95±3 |
| Another anticytokine therapy | Yes: 9 (32.1%)<br>No: 19 (67.9%) | Yes: 13 (18.6%)<br>No: 57 (81.4%) | Yes: 10 (21.3%)<br>No: 37 (78.7%) |
| Cardiovascular disease | Yes: 18 (64.3%) | Yes: 25 (35.7%) | Yes: 27 (57.4%) |

| | | | |
|---|---|---|---|
| | No: 10 (35.7%) | No: 45 (64.3%) | No: 20 (42.6%) |
| Diabetes | Yes: 6 (21.4%)<br>No: 22 (78.6%) | Yes: 9 (12.9%)<br>No: 61 (87.1%) | Yes: 13 (27.7%)<br>No: 34 (72.3%) |
| Atrial fibrillation | Yes: 4 (14.3%)<br>No: 24 (85.7%) | Yes: 4 (5.7%)<br>No: 66 (94.3%) | Yes: 4 (8.5%)<br>No: 43 (91.5%) |
| HIV | Yes: 0 (0%)<br>No: 28 (100%) | Yes: 0 (0%)<br>No: 70 (100%) | Yes: 1 (2.1%)<br>No: 46 (97.9%) |
| Ongoing immunosupressive therapy for concurrent disease | Yes: 0 (0%)<br>No: 28 (100%) | Yes: 0 (0%)<br>No: 70 (100%) | Yes: 0 (0%)<br>No: 47 (100%) |
| Chronic liver disease | Yes: 0 (0%)<br>No: 28 (100%) | Yes: 2 (2.9%)<br>No: 68 (97.1%) | Yes: 3 (6.4%)<br>No: 44 (93.6%) |
| Oncological treatment | Yes: 1 (3.6%)<br>No: 27 (96.4%) | Yes: 3 (4.3%)<br>No: 67 (95.7%) | Yes: 4 (8.5%)<br>No: 43 (91.5%) |
| Chronic respiratory disease | Yes: 4 (14.3%)<br>No: 24 (85.7%) | Yes: 4 (5.7%)<br>No: 66 (94.3%) | Yes: 8 (17%)<br>No: 39 (83%) |
| Tuberculosis | Yes: 0 (0%)<br>No: 28 (100%) | Yes: 1 (1.4%)<br>No: 69 (98.6%) | Yes: 2 (4.3%)<br>No: 45 (95.7%) |

Supplementary Table S4. Contingency tables for detected clusters

| Cluster | Group | Discharge | Death | OR, 95% CI | *p*-value | adj. *p* |
|---|---|---|---|---|---|---|
| 1 | Treatment arm | 24 | 4 | 0.51, [0.095, 2.33] | 0.5 | 1 |
| | Reference arm | 21 | 7 | | | |
| 2 | Treatment arm | 69 | 1 | 0.19, [0.004, 1.77] | 0.21 | 0.63 |
| | Reference arm | 65 | 5 | | | |
| 3 | Treatment arm | 41 | 6 | 0.32, [0.09, 0.98] | 0.046 | 0.138 |
| | Reference arm | 32 | 15 | | | |

Supplementary figure S5. Covariate balance for detected clusters.

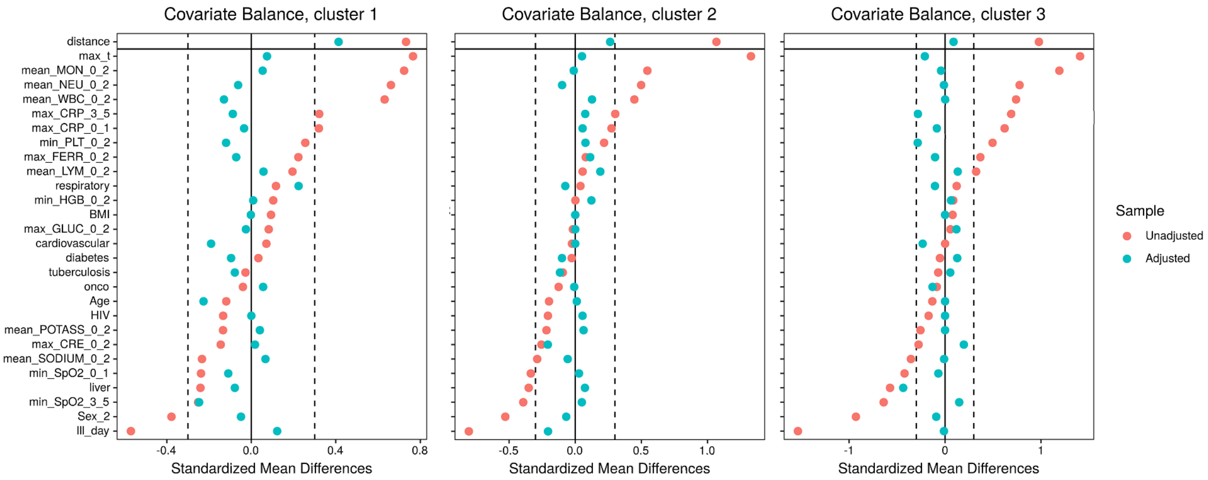


Supplementary Table S5. List of main *R* packages used in the project

| Packages | Purpose | Reference |
|---|---|---|
| ggplot, ggpubr | General data visualization. | [1-2] |
| dplyr, tidyr | Data preprocessing. | [3-4] |
| cr17 | Survival analysis | [5] |
| pheatmap | Heatmaps visualization and clustering. | [6] |
| NbClust | Choice of appropriate number of clusters. | [7] |
| MatchIt, cobalt | Propensity score matching and covariate balance visualization. | [8] |

1. H. Wickham. ggplot2: Elegant Graphics for Data Analysis. Springer-Verlag, New York, 2016.
2. Alboukadel Kassambara (2019). ggpubr: 'ggplot2' Based Publication Ready Plots. R package version 0.2.4. https://CRAN.R-project.org/package=ggpubr
3. Hadley Wickham, Romain François, Lionel Henry and Kirill Müller (2020). dplyr: A Grammar of Data Manipulation. R package version 1.0.2. https://CRAN.R-project.org/package=dplyr
4. Hadley Wickham and Lionel Henry (2019). tidyr: Tidy Messy Data. R package version 1.0.0. https://CRAN.R-project.org/package=tidyr
5. cr17: Testing Differences Between Competing Risks Models and Their Visualisations. https://CRAN.R-project.org/package=cr17
6. Raivo Kolde (2019). pheatmap: Pretty Heatmaps. R package version 1.0.12. https://CRAN.R-project.org/package=pheatmap
7. Malika Charrad, Nadia Ghazzali, Veronique Boiteau, Azam Niknafs (2014). NbClust: An R Package for Determining the Relevant Number of Clusters in a Data Set. Journal of Statistical Software, 61(6), 1-36. http://www.jstatsoft.org/v61/i06/
8. Daniel E. Ho, Kosuke Imai, Gary King, Elizabeth A. Stuart (2011). MatchIt: Nonparametric Preprocessing for Parametric Causal Inference. Journal of Statistical Software, Vol. 42, No. 8, pp. 1-28. URL https://www.jstatsoft.org/v42/i08/

Supplementary figure S6. Subgroup analysis according to the physical characteristics

| Subgroups | Reference Alive | Reference Death | Treatment Alive | Treatment Death | Odds Ratio [95% CI] |
|---|---|---|---|---|---|
| ***Sex*** | | | | | |
| Sex Male | 72 | 7 | 72 | 8 | 1.14 [0.39, 3.32] |
| Sex Female | 55 | 12 | 60 | 6 | 0.46 [0.16, 1.30] |
| ***SpO2*** | | | | | |
| min_SpO2_3_5 >=94 | 81 | 7 | 79 | 6 | 0.88 [0.28, 2.73] |
| min_SpO2_3_5 <94 | 46 | 12 | 53 | 8 | 0.58 [0.22, 1.54] |
| min_SpO2_0_1 >=94 | 68 | 8 | 79 | 7 | 0.75 [0.26, 2.18] |
| min_SpO2_0_1 <94 | 59 | 11 | 53 | 7 | 0.71 [0.26, 1.96] |
| ***Temperature*** | | | | | |
| max_t >=38 | 64 | 12 | 80 | 5 | 0.33 [0.11, 1.00] |
| max_t <38 | 63 | 7 | 52 | 9 | 1.56 [0.54, 4.47] |
| ***Day of illness*** | | | | | |
| Ill_day >=7 | 80 | 13 | 79 | 6 | 0.47 [0.17, 1.29] |
| Ill_day <7 | 47 | 6 | 53 | 8 | 1.18 [0.38, 3.66] |
| ***BMI*** | | | | | |
| BMI >=30 | 56 | 8 | 69 | 5 | 0.51 [0.16, 1.64] |
| BMI <30 | 71 | 11 | 63 | 9 | 0.92 [0.36, 2.37] |
| ***Age*** | | | | | |
| Age >=60 | 60 | 16 | 61 | 10 | 0.61 [0.26, 1.46] |
| Age <60 | 67 | 3 | 71 | 4 | 1.26 [0.27, 5.83] |

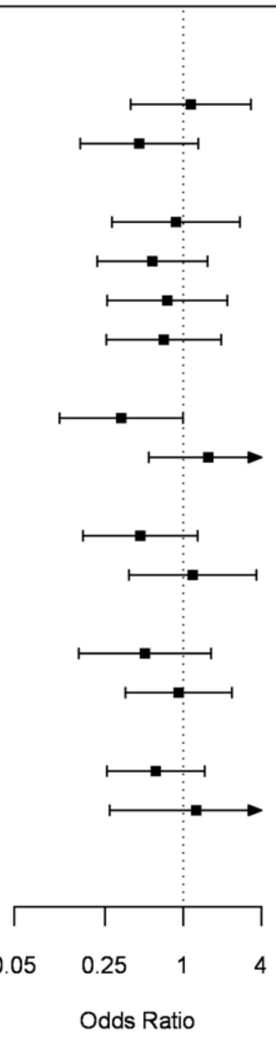

Supplementary figure S7. Subgroup analysis according to the admission biochemistry parameters

| Subgroups | Reference Alive | Reference Death | Treatment Alive | Treatment Death | Odds Ratio [95% CI] |
|---|---|---|---|---|---|
| ***Sodium*** | | | | | |
| mean_SODIUM_0_2 >=136 | 104 | 16 | 104 | 13 | 0.81 [0.37, 1.77] |
| mean_SODIUM_0_2 <136 | 23 | 3 | 28 | 1 | 0.27 [0.03, 2.81] |
| ***Potassium*** | | | | | |
| mean_POTASS_0_2 >=3.5 | 118 | 17 | 121 | 9 | 0.52 [0.22, 1.20] |
| mean_POTASS_0_2 <3.5 | 9 | 2 | 11 | 5 | 2.05 [0.32, 13.16] |
| ***Glucose*** | | | | | |
| max_GLUC_0_2 >=6.8 | 63 | 16 | 59 | 11 | 0.73 [0.32, 1.71] |
| max_GLUC_0_2 <6.8 | 64 | 3 | 73 | 3 | 0.88 [0.17, 4.50] |
| ***Ferritin*** | | | | | |
| max_FERR_0_2 >=490 | 59 | 14 | 64 | 10 | 0.66 [0.27, 1.60] |
| max_FERR_0_2 <490 | 68 | 5 | 68 | 4 | 0.80 [0.21, 3.11] |
| ***C-reactive protein*** | | | | | |
| max_CRP_3_5 >=100 | 47 | 18 | 48 | 9 | 0.49 [0.20, 1.20] |
| max_CRP_3_5 <100 | 80 | 1 | 84 | 5 | 4.76 [0.54, 41.65] |
| max_CRP_0_1 >=100 | 48 | 16 | 47 | 10 | 0.64 [0.26, 1.55] |
| max_CRP_0_1 <100 | 79 | 3 | 85 | 4 | 1.24 [0.27, 5.71] |
| ***Creatinine*** | | | | | |
| max_CRE_0_2 >=0.115 | 16 | 10 | 27 | 6 | 0.36 [0.11, 1.16] |
| max_CRE_0_2 <0.115 | 111 | 9 | 105 | 8 | 0.94 [0.35, 2.53] |

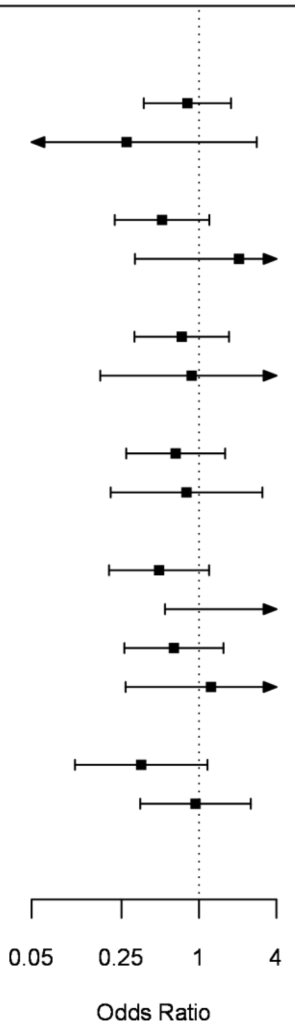

Supplementary figure S7. Subgroup analysis according to the co-morbidities.

| Subgroups | Reference Alive | Reference Death | Treatment Alive | Treatment Death | Odds Ratio [95% CI] |
|---|---|---|---|---|---|
| ***Tuberculosis*** | | | | | |
| tuberculosis Yes | 4 | 0 | 3 | 0 | 1.29 [0.02, 82.49] |
| tuberculosis No | 123 | 19 | 129 | 14 | 0.70 [0.34, 1.46] |
| ***Respiratory*** | | | | | |
| respiratory Yes | 11 | 3 | 14 | 3 | 0.79 [0.13, 4.68] |
| respiratory No | 116 | 16 | 118 | 11 | 0.68 [0.30, 1.52] |
| ***Oncological*** | | | | | |
| onco Yes | 9 | 1 | 6 | 3 | 4.50 [0.37, 54.16] |
| onco No | 118 | 18 | 126 | 11 | 0.57 [0.26, 1.26] |
| ***Liver*** | | | | | |
| liver Yes | 9 | 0 | 5 | 0 | 1.73 [0.03, 99.95] |
| liver No | 118 | 19 | 127 | 14 | 0.68 [0.33, 1.43] |
| ***HIV*** | | | | | |
| HIV Yes | 1 | 0 | 1 | 0 | 1.00 [0.01, 92.42] |
| HIV No | 126 | 19 | 131 | 14 | 0.71 [0.34, 1.47] |
| ***Diabetes*** | | | | | |
| diabetes Yes | 33 | 2 | 26 | 3 | 1.90 [0.30, 12.25] |
| diabetes No | 94 | 17 | 106 | 11 | 0.57 [0.26, 1.29] |
| ***Cardivascular*** | | | | | |
| cardiovascular Yes | 67 | 8 | 62 | 11 | 1.49 [0.56, 3.94] |
| cardiovascular No | 60 | 11 | 70 | 3 | 0.23 [0.06, 0.88] |

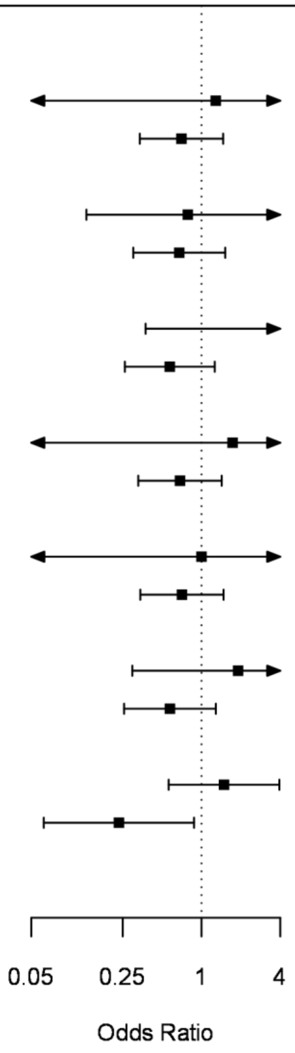

Supplementary table S6. Comparison of ruxolitinib and dexamethasone across COVID-19 severity based on computer tomography evaluation. Odds ratio below 1.0 is in favor of ruxolitinib, above 1.0 – in favor of dexamethasone.

| Computer tomography stage | Odds ratio (OR) | OR 95% CI | P-value |
|---|---|---|---|
| Stage 1 (<25%) | N/A (no cases of mortality) | N/A (no cases of mortality) | 1.0 |
| Stage 2 (25-50%) | 0.32 | 0.17-1.9 | 0.56 |
| Stage 3 (51-75%) | 0.59 | 0.37-2.13 | 0.59 |

Supplementary table S7. Incidence of hematological adverse events in the ruxolitinib (treatment) and dexametasone (reference) arms.

| Test | Group | 0 | 1 | 2 | 3 | 4 | *p* |
|---|---|---|---|---|---|---|---|
| HGB | Reference | 56 (38.4%) | 67 (45.9%) | 15 (10.3%) | 7 (4.8%) | 1 (0.7%) | 0.69 |
| | Treatment | 50 (34.2%) | 71 (48.6%) | 20 (13.7%) | 5 (3.4%) | 0 (0%) | |
| LYM | Reference | 42 (28.8%) | 46 (31.5%) | 32 (21.9%) | 26 (17.8%) | 0 (0%) | 0.15 |
| | Treatment | 27 (18.5%) | 46 (31.5%) | 42 (28.8%) | 29 (19.9%) | 2 (1.4%) | |
| PLT | Reference | 107 (73.3%) | 34 (23.3%) | 4 (2.7%) | 1 (0.7%) | 0 (0%) | 0.042 |
| | Treatment | 86 (58.9%) | 54 (37%) | 4 (2.7%) | 1 (0.7%) | 1 (0.7%) | |
| WBC | Reference | 109 (74.7%) | 24 (16.4%) | 10 (6.8%) | 3 (2.1%) | 0 (0%) | 0.33 |
| | Treatment | 94 (64.4%) | 32 (21.9%) | 14 (9.6%) | 5 (3.4%) | 1 (0.7%) | |

Supplementary table S8. Rare adverse events and their incidence

| Ruxolitnib arm | Incidence, N(%) | Dexamethasone arm | Incidence, N(%) |
|---|---|---|---|
| Oral candidiasis | 2 (1.4%) | Acute kidney injury | 2 (1.4%) |
| Herpes Zoster | 1 (0.7%) | Purulent cholecystitis | 1 (0.7%) |
| Toxic hepatitis grade 3 | 1 (0.7%) | Purulent otitis | 1 (0.7%) |
| Gastrointestinal bleeding | 1 (0.7%) | Acute pancreatitis | 1 (0.7%) |
| Spontaneous intra-abdominal hemorrhage | 1 (0.7%) | | |
